# Effectiveness of the mRNA BNT162b2 vaccine against SARS-CoV-2 severe infections in the Israeli over 60 population: a temporal analysis done by using the national surveillance data

**DOI:** 10.1101/2021.09.27.21264130

**Authors:** Stefano De Leo

## Abstract

Last August, when the *delta* variant became the dominant infection strain, Israel, one of the countries with the highest levels of vaccination in the world, faced a scary pandemic wave. The frighteningly increasing number of infections was seen as *the perfect storm* to test the effectiveness of the mRNA BNT162b2 vaccine. The new surge forced the government to use a booster shot to protect the most vulnerable age groups. Starting from the August national surveillance data, we analysed the *temporal* effectiveness of vaccination against severe infections in the Israeli over 60 population. The study shows that the two-dose vaccine still works in preventing people from getting seriously sick but not with the same effectiveness observed in the first months of 2021. However, the observed temporal increase of the vaccine effectiveness in Israel, during August, suggests a correlation with the increase of the population protected by the booster shot.

---

With the change over time of the virus and the arrival of new pandemic waves, it is important to constantly compare active cases and severe infections within the vaccinated and unvaccinated population. The monitoring of the temporal vaccine effectiveness plays a fundamental role in guiding individual choices and decisions of the local authorities. From January 24 to April 3, a deep and detailed look into the Israeli fully (seven day or longer the second dose) vaccinated and unvaccinated groups showed, in the over 16 Israeli population, an effectiveness of the mRNA BNT162b2 (Pfizer-BioNTech proprietary name) vaccine of 97.5% (97.1%-97.8%) against severe infections [1]. During the 70 days of this analysis, there were 232268 Sars-Cov-2 confirmed cases, with 3.31% hospitalisations, 1.93% sever infections, and 0.48% deaths. The prevalence of the B.1.1.7, also known as *alpha* [2], variant among the Sars-Cov-2 infections was of 94.5%. So, in the first months after the two doses vaccine administration and in the presence of a prevalent *α* variant scenario, the BNT162b2 vaccine was highly effective against severe infections across all age groups. Classified with the acronym B.1.617.2, the *delta* [2] variant, which is nowdays dominant throughout the planet, was responsible, during August, for a dramatic surge of new infections in Israel with a peak of a seven-day average greater than 10 thousand new daily confirmed cases approaching the number of the Israeli peak in the past winter. This implies almost 1100 new confirmed cases per million of inhabitants and this represents one of the world’s highest daily infection rates. Notwithstanding the fact that Israel was the first country in the World to vaccinate the almost totality of its citizens against Srars-Cov-2, the virus nightmare came back. Why did this happend? In Israel, by June, all restrictions, including indoor masking, were abolished and this together with the decrease in vaccine protection, expected around six months after receiving the second shot [4], and the high contagiousness of the *delta* variant, combined with its very powerful viral load, surprised Israel creating the perfect storm and forced the health authorities to approve at the end of July the administration of a third (booster) dose of the BNT162b2 vaccine for the over 60 population who had received a second dose of vaccine at least 5 months earlier.

In almost all countries of the world, the vaccination campaign was initially characterized by a demand that exceeded the daily national supply and, then, followed by a vaccine hesitancy caused by a lack of knowledge or, more seriously, by disinformation and scepticism propagated in social networks. Covid-19 vaccinations is optional worldwide rather than mandatory, with very few exceptions. In many countries, the government authorities have introduced measures to stimulate vaccination by allowing entry in bars, restaurants, swimming pools, gyms, tourist attractions, cultural or sport events only for the *green pass* holders. Admission to essential services for the population, see for example hospitals, clinics, pharmacies, supermarket, and so on, was, obviously, not restricted. The green pass requirement is surely a tool to stimulate vaccination but *not* only this. Indeed, in view of the real world effectiveness results, it also represents an important tool to allow the reopening of the economy by *limiting* infections and, consequently, avoiding the overcrowding of the health system. At this particular time, when a small, but yet important, part of the population has to be vaccinated, a careful reading of the data, coming from the national surveillance systems, is of fundamental importance, not only to understand the effectiveness of vaccination, but also to correctly communicate with the people. Misinterpreted data can lead to equivocal conclusions and arouse doubts and distrust in the population slowing down the vaccination campaign.

In this letter, individuals were defined as unvaccinated if they had not received any dose of the Pfizer-BioNTech (BNT162b2 international non-proprietary name) and as fully vaccinated if they had received at least two doses of the vaccine. At the day September 2, in Israel, the third booster shot campaign additionally protected almost 40% of the population that received two doses of BNT162b2 [6]. We aim to show how a simple but careful analysis of the surveillance data allows to obtain information about the *temporal* effectiveness of the vaccine and consequently, if necessary, to address local authorities to face new pandemic waves. On September 2, according to the surveillance data by the Israel Ministry of Health, daily updated in [6], both the active cases for the over 30 population and the severe infections for the over 70 population were greater in the fully vaccinated group than in the unvaccinated one, see Table 1(a). For example by looking at severe infections, we find 82 cases in the 70-79 age fully vaccinated group against the 74 of the unvacinated one, 60 in the fully vaccinated 80-89 age group against the 34 of the unvaccinated one, and, finally, in the over 90 population, we observe an even number of severe infections, i.e. 20. To simplify our presentation, we will avoid considering cases with still incomplete vaccination, the data corresponding at such a group are available in [6].

**Table 1:**
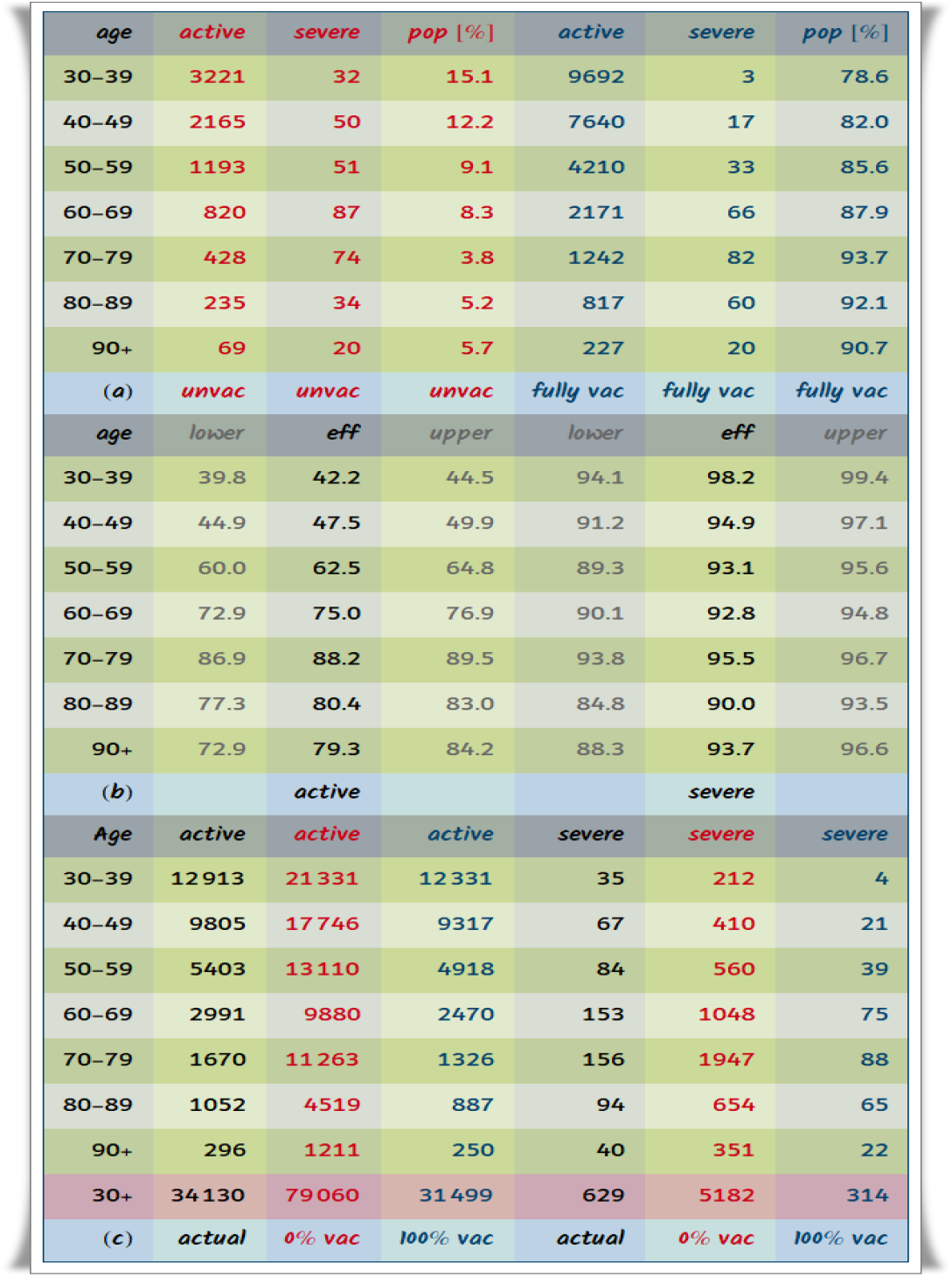
The Israeli surveillance data of day September 2. Active cases and severe infections are reported for different age groups in (a), where we also find the percentage of the unvaccinated and fully vaccinated population. By using these data, we can calculate the mRNA BNT162b2 vaccine effectiveness and its 95% confidence interval for active cases and severe infections, the results are given in (b). Finally in (c), we simulate two limit situations: the one in which all the population is fully vaccinated and the one in which it is completely unvaccinated. In the last case, for the over 30 population, we would go from 34130 to 79060 active cases and from 629 to 5182 severe infections. This means that, without any vaccination and in the same circumstances of reopening, we should have an increase of 232% of active cases and of 824% of severe infections.

Before jumping to *wrong* conclusions, let’s look at the fourth and seventh column of the Table 1(a), where the percentage of unvaccinated and fully vaccinated population is reported. These percentages allow us to simulate what would have happened in two extreme cases, i.e. the case of the lack of vaccine and the case of an entire population which is fully vaccinated. It is indeed more understandable to a wider audience to present sample statistics as estimates of outcomes that would be obtained if the total population were unvaccinated or fully vaccinated. For example, in the last case, from the Israeli data, we would have found, in the 70-79 age group, 82 × 100/93.7≈ 88 severe infections, in the 80-89 one, 60 × 100/92.1 ≈ 65, and, finally, in the over 90 population, 20 × 100/90.7 ≈ 22 severe infections. Repeating such a simulation, now supposing that the entire population was unvaccinated, we obtain, in the 70-79 age group, 74 × 100/3.8. ≈1947 severe infections, in the 80-89 age group, 34 × 100/5.2 ≈ 654, and, finally, in the over 90 population, 20 × 100/5.7 ≈ 351. In this simplified presentation, the numbers of *normalised* severe infections shed light on the real situation and give us a clearer view of the effectiveness of the vaccine. For example, the 88 severe infections, found in the 70-79 age group supposing that the entire population was fully vaccinated, represent a reduction of 1859 severe infections with respect to the 1947 that we would have found if we hadn’t had the vaccine available. This corresponds to an effectiveness of 1859/1947 ≈ 95.5%. By using the same technique, in the 80-89 and over 90 age groups, we observe an effectiveness of 589/654 ≈ 90.1% and 329/351 ≈ 93.7%, respectively.

In Table 1(b), for each age group, we give the vaccine effectiveness calculated by using the *relative risk* [7]

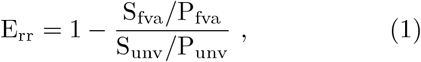

where S is the number of severe infections, P the population, and the lower scripts refer to the fully vaccinated (fva) and unvaccinated (unv) population. In this Table, we also find the lower and upper limits of the 95% confidence interval, calculated by using

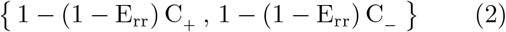

where

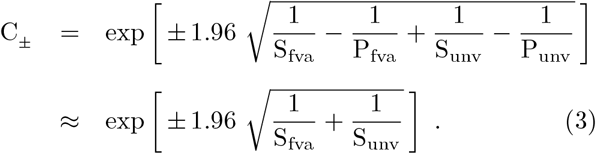

The confidence interval represents the range of values that are considered to be plausible for our outcomes, in this particular case the plausible interval of the vaccine effectiveness. As it can be seen from the previous formula, the width of a confidence interval depends on the sample size. From Table 1(b), for active cases, we see that the smallest interval belongs to the the 30-39 age group. This is expected due to the fact that the younger part of the population is more exposed to the contagion than the older one. For severe infections, the different gravity in each age group acts as a normalisation and, consequently, we find a same width of the 95% confidence bands.

For active cases, we observe the lowest effectiveness in the 30-39 age range, 42.2%, and the highest one in the 70-79 age range, 88.2%. For severe infections, a very high effectiveness is found in all the age ranges: 98.2% (30-39), 94.9% (40-49), 93.1% (50-59), 92.8% (60-69), 95.5% (70-79), 90.0% (80-89), 93.7% (over 90). Let us now briefly discuss the overall effectiveness in the over 30 population. From Table 1(c), for active cases, we find an effectiveness of (79060 − 31499)/79060 ≈ 60.2% and, for severe infections, an effectiveness of (5182 − 314)/5182 ≈ 93.9%. It is important to recall that this analysis refers to the day September 2, so after the application of the booster shot in part of the vulnerable age groups. Observe, that an active over 30 cases effectiveness of 60.2% clearly shows, as anticipated in our introduction, that the *green pass* requirement is *not* only a measure to stimulate vaccination but also an important tool to reduce infections and protect people. This obviously does not mean that other containment measures, such as wearing masks, social distancing and infection screening, can be completely forgotten. Vaccination, screening and containment policy, indeed all of them, are important to gradually return to normality. Obviously, a full normality can be only reached when a large scale vaccine immunization in the worldwide population will be achieved.

Let’s take a step back in time and see what happened in Israel during the month of August. In Table 2, we find the daily data of severe infections in the over 60 population during the period from August 3 to September 1. The absolute numbers of severe infections are greater in the fully vaccinated population than in the unvaccinated one, see Table 2 or Figure 1(a). This is as observed before, absolutely expected when when we carry out a successful vaccination campaign as the one done by Israel where almost 90% of the over 60 population was fully vaccinated. In the Table 2, we also find the severe infections per hundred thousand inhabitants both for the fully vaccinated and for the unvaccinated group. From these data, we can estimate the situation in two opposite cases: the first case the one in which the entire population is unvaccinated and the second case the one in which the whole population is fully vaccinated, see Figure 1(b). The plots show the dramatic situation that would have occurred in the case of the lack of vaccines. Indeed, by looking at Figure 1 (a) and (b), we observe an increase of a factor 10 in severe infections. This scary increase would have generated a serious crisis in the Israeli health system.

**Table 2:** The Israeli surveillance data for severe infections in the over 60 population from the day August 3 to the day September 1. For each day, we find the absolute numbers of severe infections in the unvaccinated (red color) and fully vaccinated (blue color) groups. To calculate the vaccine effectiveness, the absolute numbers have to be normalised to the respective population. The numbers of severe infections per hundred thousand inhabitants appear for the unvaccinated group in the fourth and ninth column and for the fully vaccinated one in the fifth and tenth column. The vaccine effectiveness pass from (45.2 − 10.8)/45.2 ≈ 76.8% of the day August 3 to (279.2 − 17.4)/279.2 ≈ 93.8%.

| date | severe | severe | sev/100K | sev/100K | date | severe | severe | sev/100K | sev/100K |
| --- | --- | --- | --- | --- | --- | --- | --- | --- | --- |
| 8/03 | 42 | 141 | 45.2 | 10.8 | 8/18 | 158 | 271 | 185.6 | 20.5 |
| 8/04 | 44 | 141 | 47.6 | 10.7 | 8/19 | 154 | 271 | 182.4 | 20.5 |
| 8/05 | 51 | 156 | 55.5 | 11.9 | 8/20 | 166 | 266 | 197.5 | 20.1 |
| 8/06 | 52 | 163 | 56.8 | 12.4 | 8/21 | 184 | 279 | 219.7 | 21.1 |
| 8/07 | 63 | 193 | 68.9 | 14.7 | 8/22 | 181 | 297 | 217.9 | 22.5 |
| 8/08 | 76 | 211 | 83.6 | 16.1 | 8/23 | 195 | 280 | 236.6 | 21.2 |
| 8/09 | 84 | 213 | 93.0 | 16.2 | 8/24 | 199 | 272 | 243.5 | 20.5 |
| 8/10 | 87 | 219 | 96.9 | 16.7 | 8/25 | 214 | 267 | 263.8 | 20.2 |
| 8/11 | 88 | 223 | 98.7 | 17.0 | 8/26 | 214 | 255 | 265.8 | 19.2 |
| 8/12 | 98 | 252 | 110.7 | 19.2 | 8/27 | 216 | 255 | 269.4 | 19.2 |
| 8/13 | 108 | 255 | 122.5 | 19.4 | 8/28 | 213 | 261 | 266.6 | 19.7 |
| 8/14 | 117 | 271 | 133.3 | 20.6 | 8/29 | 238 | 271 | 300.1 | 20.4 |
| 8/15 | 125 | 280 | 143.4 | 21.3 | 8/30 | 234 | 263 | 296.6 | 19.8 |
| 8/16 | 134 | 262 | 155.0 | 19.9 | 8/31 | 219 | 244 | 280.4 | 18.4 |
| 8/17 | 147 | 279 | 171.4 | 21.2 | 9/01 | 217 | 231 | 279.2 | 17.4 |
|  | unvac | full vac | unvac | full vac |  | unvac | fully vac | unvac | fully vac |

**Figure 1:**
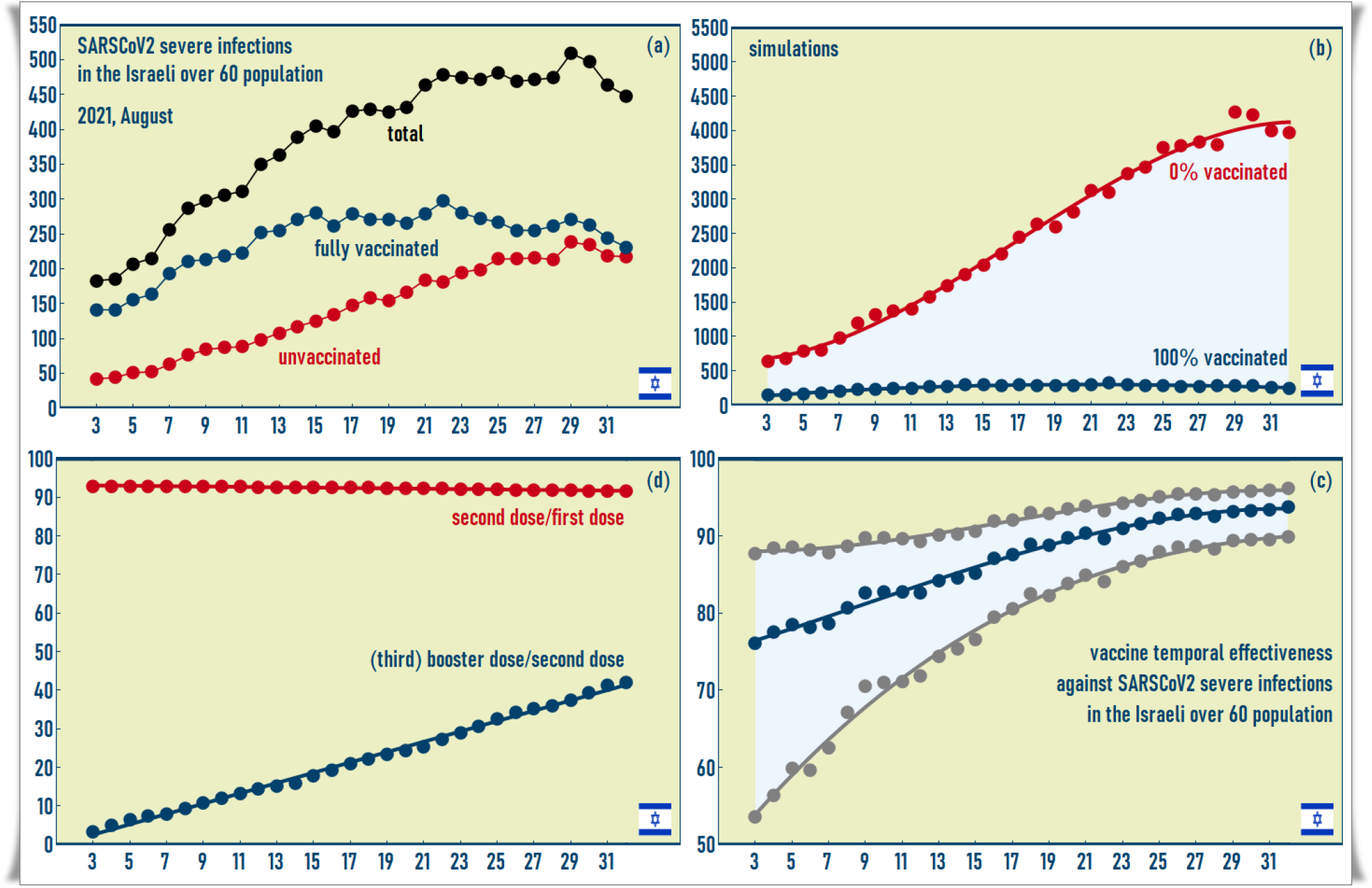
In (a), the total number of patients in the population over 60 with severe infection is plotted with a black line. In the same graph, we also find the number of severely infected patients in the unvaccinated group (red line) and fully vaccinated (blue line) groups. The simulations of two extreme cases i.e. that of a 100% unvaccinated population and that of a 100% fully vaccinated population are shown in (b). By comparing the plots in (a) and in (b), we easily conclude that, in the absence of a vaccine, infections would have increased by a factor of 10. Starting from the national surveillance data, it is possible to calculate the vaccine effectiveness for each day, see (c). The cubic curve, obtained by a linear regression, clearly shows a temporal increase of the effectiveness in correspondence to the application of the booster dose, see (d).

Let us now study, in detail, the data from Table 2. We begin by comparing the first day of our analysis with the last one. At August 3, in the unvaccinated group, we find 45.2 severe infections per hundred thousand inhabitants while, in the fully vaccinated population, 10.8. This means an effectiveness of (45.2 − 10.8)/45.2 ≈ 76.1%. This is in perfect agreement with the result recently reported in [8] showing that the Pfizer-BioNTech vaccine effectiveness wanes after 120 day and the protection drops to about 77%. At September 1, we have 279.2 severe infections in the unvaccinated group against the 17.4 of the fully vaccinated one. This implies an effectiveness of 261.8/279.2 ≈ 93.8%, recovering the initial effectiveness of the Pfizer-BioNTech vaccine. Figure 1(c) shows the *temporal* effectiveness observed during August with its lower and upper 95% confidence bounds. The decrease in time of the 95% confidence interval is due to the fact that, in August, Israel faced a new pandemic wave with an increasing number of severe infections. This has the statistical effect to reduce the error on the effectiveness. For example, in the unvaccinated population, we pass from 45.2 severe infections per hundred thousand inhabitants of the day August 3 to 279.2 on September 1, this represents a factor 6.2, and, in the fully vaccinated group, from 10.8 to 17.4, i.e. a factor 1.6. These factors are also important because they show how the pandemic has a different growth within the unvaccianted and the fully vaccinated group, see Figure 1(a). This pandemic ratio can be considered as an alternative way to see the effectiveness of the vaccination in a country. Once the behaviour of the confidence interval and the different growth between unvaccinated and fully vaccinated population have been clarified, the increase in the vaccine effectiveness still remains to be explained.

By using a linear regression, we modelled the temporal effectiveness curve, see Figure 1 (c), and the percentage of the fully vaccinated population that was protected by a third booster dose, see the blue curve in Figure 1 (d), by a cubic

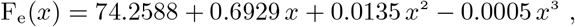

and a linear polynomial

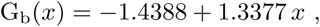

respectively. From these equations, we can extract important information about the effect of the booster shoot in protecting the Israeli population. In the first fifteen days of August the effectiveness of the vaccine shows a linear increase with an angular coefficient of about 0.69 which is almost half of the angular coefficient that characterizes the temporal course of vaccination with a booster dose, i.e. 1.34. This can also be see graphically by looking at Figures (c) and (d). The straight line which approximate the effectiveness in the first days of August, see in (c), and the straight line representing the booster shot trend are parallel. By observing that Figure (c) and (b) having a factor of two of difference in their ordinate axis, we graphically found the factor two mentioned above. The fact that the two straight lines seem to join is instead a pure coincidence due to the fact that at the end of August the Israeli authorities had vaccinated with the booster dose almost 40% of the two-dose vaccinated population and that the vaccine effectiveness at the beginning of August was about 80%.

The *golden rule* of our analysis is that, at the beginning of the booster immunization, for each 2 *p*% of the population protected by the booster dose, we find an increase of *p*% in the vaccine effectiveness. The cubic nature of the temporal behaviour of the effectiveness also allows to determine the day at which the quadratic and cubic term act as a brake and tend to stabilize the effectiveness of the vaccine,

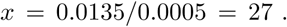

This letter, whose spirit is mainly didactic and informative, has the objective of clarifying how to read the national surveillance data and to draw indicative conclusions on the vaccine effectiveness. By using the Israeli data, referring to the month of August, we have seen how the decrease in the vaccine effectiveness, caused by the time elapsed from the application of the second dose and the advent of a new pandemic wave with the *delta* variant as dominant infection strain, was *correctly* and *promptly* addressed by the local authorities by using a (third) booster dose for the Israeli population. This pandemic war will only be won when all the people in the world will gain and use the opportunity to vaccinate. Waiting for this moment, a constant analysis of pandemic data done by the scientific community and its disclosure for a wide audience will play a fundamental to gain decisive battles against fake news and disinformation.

This work is dedicated to Professor *Erasmo Recami*, recently passed away. His enthusiasm, his preparation, his passion and his continuous words of encouragement will always be remembered with love and gratitude.

**QR code for 10.1101/2021.09.27.21264130**

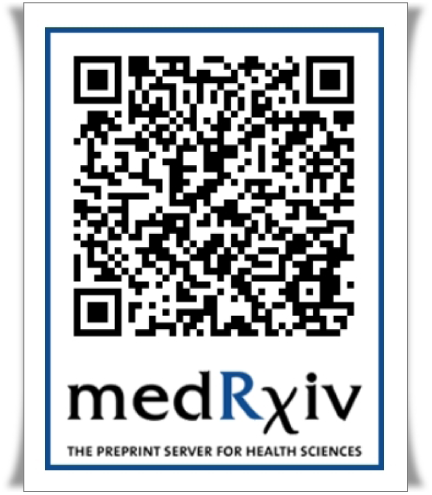

## Data Availability

Data are available in the Tables of the paper.

https://datadashboard.health.gov.il/COVID-19/

